# On-scalp magnetoencephalography based on optically pumped magnetometers to investigate temporal lobe epilepsy

**DOI:** 10.1101/2024.10.21.24315793

**Authors:** Odile Feys, Vincent Wens, Chantal Depondt, Estelle Rikir, Nicolas Gaspard, Wim Van Paesschen, Alec Aeby, Olivier Bodart, Evelien Carrette, Niall Holmes, Matthew Brookes, Maxime Ferez, Pierre Corvilain, Xavier De Tiège

**Affiliations:** Université libre de Bruxelles (ULB), Hôpital Universitaire de Bruxelles (HUB), Hôpital Erasme, Department of neurology, Bruxelles, Belgium; Université libre de Bruxelles (ULB), ULB Neuroscience Institute (UNI), Laboratoire de neuroanatomie et neuroimagerie translationnelles (LN2T), Bruxelles, Belgium; Université libre de Bruxelles (ULB), Hôpital Universitaire de Bruxelles (HUB), Hôpital Erasme, Department of translational neuroimaging, Bruxelles, Belgium; Yale University, Department of neurology, New Haven, CT, USA; Katholieke Universiteit Leuven (KULeuven), Universitair Ziekenhuis Leuven (UZ Leuven), Department of neurology, Leuven, Belgium; Université libre de Bruxelles (ULB), Hôpital Universitaire de Bruxelles (HUB), Hôpital universitaire des enfants Reine Fabiola (HUDERF), Department of pediatric neurology, Bruxelles, Belgium; Université de Liège (ULiège), Centre hospitalier universitaire de Liège (CHU Liège), Department of neurology, Liège, Belgium; Universiteit Gent, Universitair Ziekenhuis Gent (UZ Gent), Department of neurology, Gent, Belgium; University of Nottingham, School of Physics and Astronomy, Sir Peter Mansfield Imaging Centre, Nottingham, United Kingdom

## Abstract

Cryogenic magnetoencephalography (MEG) has a lower yield in temporal lobe epilepsy (TLE) than in extra-TLE (ETLE). The advent of optically pumped magnetometers (OPMs) might change this thanks to on-scalp MEG, which allows sensors to be placed closer to the brain and the design of bespoke sensor arrays to target specific brain regions. This study aims to investigate the detection and localization accuracy of interictal epileptiform discharges (IEDs) using on-scalp MEG in TLE and the added-value of face-OPMs for temporal IED detection/localization.

Eleven patients underwent a 1-h MEG recording with OPMs placed both on the scalp (flexible cap, scalp-OPMs) and on the face (3D-printed glass-like structure, face-OPMs). Nine patients also underwent cryogenic MEG. IEDs were visually detected, averaged and localized using distributed source reconstruction. On-scalp MEG IED amplitude and signal-to-noise (SNR) were assessed and compared with cryogenic MEG when more than 10 IEDs were detected. Neural sources with and without face-OPMs were compared. The correlation between face- and scalp-OPMs was assessed.

A mean of 13 IEDs/patient was detected using on-scalp MEG (mean amplitude: 3.3pT, mean SNR: 9.4) and localized in the (medial, anterior, basal, lateral or posterior) temporal lobe. Three patients had >10 IEDs in on-scalp and cryogenic MEG signals with amplitude and SNR that were either higher or similar for the on-scalp MEG recording compared with cryogenic MEG, and sources were separated by 8-11 mm. In two other patients, on-scalp MEG source locations were confirmed by gold-standard methods (surgical resection cavity, n=1; stereo-electroencephalography, n=1). Face-OPMs had a clear added-value (i.e., IED detection and localization) in one patient with antero-medial TLE. Face-OPM signals were correlated with scalp-OPM signals in most patients, showing that face-OPMs recorded brain activity.

This study shows that on-scalp MEG is able to detect and localize IEDs in TLE and to discriminate irritative zones from different key (medial, anterior, basal, lateral or postero-lateral) temporal areas with similar or enhanced SNR than cryogenic MEG. Face-OPMs have a clear added-value in patients with anterior/medial TLE and increase the spatial coverage of the temporal lobe. This study paves the way for the future use of on-scalp MEG in patients with refractory TLE or with other brain disorders affecting the temporal lobe such as, e.g., Alzheimer’s disease.

## Introduction

Cryogenic magnetoencephalography (MEG) has demonstrated its clinical value in the presurgical assessment of refractory focal epilepsy (RFE), but with a lower yield in temporal lobe epilepsy (TLE) compared with extratemporal lobe epilepsy (ETLE) (1,2). This is partly because brain magnetic field amplitude decreases with the square of brain-to-sensor distance and due to the low coverage of the temporal lobe leading to partial pick-up of the magnetic field pattern (3). The localization accuracy of cryogenic MEG for medial temporal interictal epileptiform discharges (IEDs) is thus limited due to their deep onset that reduces their detection sensitivity and to rapid anterior or latero-posterior temporal neocortical propagation pathways that are more easily picked-up by MEG sensors (4,5).

Cryogenic cooling requires MEG sensors to be housed in an adult-sized rigid helmet with a thermally insulated space that negatively impacts MEG signal-to-noise ratio (SNR; especially in medial TLE) (6) and complicates MEG recordings in non-compliant or low head circumference patients (7). The advent of miniaturized optically pumped magnetometers (OPMs) paved the way to the development of on-scalp MEG (8) with a possible increase in SNR due to reduced brain-to-sensor distance (9) and a higher lifespan/movement compliance (10,11). Case-series demonstrated that on-scalp MEG is able to detect and localize IEDs with similar or increased SNR compared with cryogenic MEG (9,12,13). Furthermore, simultaneous on-scalp MEG and stereo-electroencephalography (SEEG) recordings ultimately demonstrated that on-scalp MEG is able to accurately detect and localize IEDs from medial temporal structures (14) similarly to cryogenic MEG (15). Still, the demonstration of the ability of on-scalp MEG to record IEDs from medial temporal lobe is limited to one case (14) and requires further confirmation.

A clear strength of on-scalp MEG is that it allows the design of bespoke sensor arrays tailored to target specific brain regions with high sensor density or specific OPM locations to improve sensitivity (16). For example, mouth OPMs enhanced on-scalp MEG sensitivity to task-based hippocampal theta-band neural oscillations (17). Mouth OPMs do not conceptually differ from sphenoidal electroencephalography (EEG) electrodes that were historically used to increase the detection sensitivity of mesial temporal epileptiform discharges (18). Similarly, high-density (256 electrodes) EEG involves electrodes that cover the face to increase sensitivity to certain neural sources (19). Further, theoretical considerations on the physics of neuromagnetism demonstrated the benefits of increasing sensor coverage area from a purely scalp coverage by increasing the field-of-view of MEG and making MEG more sensitive to neuromagnetic fields (20). By bringing sensors closer to the brain and allowing the placement of additional OPMs on the face, on-scalp MEG could thus increase the yield of MEG in TLE.

This study thus aimed at (i) investigating the ability of on-scalp MEG based on OPMs to detect and localize IEDs in eleven patients with TLE, (ii) assessing the added-value of OPMs on the face for temporal IED detection, and (iii) compared the SNR of temporal IED between on-scalp MEG and cryogenic MEG.

## Methods Patients

Eleven patients with TLE underwent a 1-h on-scalp MEG recording based on the following inclusion criteria: (i) clinical follow-up in a tertiary university hospital, (ii) occurrence of IEDs originating from the temporal lobe during previous clinical short-term EEG, 24-h video-EEG or clinical MEG, (iii) ability to remain still for a 1-h recording (adults and children > 6-years-old), and (iv) written informed consent from the patient and legal representative for children. Patients optionally underwent a 1-h cryogenic MEG recording when not done for clinical purpose.

This study was approved by the institutional ethics committee from CUB Hôpital Erasme (P2019/426).

## Data acquisition

On-scalp MEG was performed using a combination of both bi-axial and tri-axial OPMs (2^nd^ and 3^rd^ generation (QuSpin Inc, USA), with a number depending on technical availability; gain: 2.7 V/nT, signal fed to a digital acquisition unit (National Instruments), sampling rate: 1,200Hz, no band-pass filter) placed on the scalp using a flexible EEG-like cap (EasyCap GmbH, Germany) on which home-made 3D- printed OPM sensor holders were sewn. OPMs were preferentially positioned to cover the temporal lobes according to the conventional 10-10 placement, and at additional positions around the ears on the flexible EEG-like cap (Figure 1). Additionally, following the framework of mouth OPMs and sphenoidal EEG electrodes, four OPMs were positioned at level of the maxillary sinuses (henceforth referred to as “face-OPMs” in contrast with “scalp-OPMs” for those positioned on the scalp, still we shall continue to refer to "on-scalp MEG" as the OPM montage that includes both face-OPMs and scalp-OPMs) using a 3D-printed homemade designed glasses-like structure (Figure 1). These face- OPMs were used to maximize patients’ comfort (compared with mouth-OPMs) and increase the sensitivity to the spatial characteristics of hippocampal magnetic fields (17). On-scalp MEG recordings took place in a dedicated magnetically shielded room (MSR; Compact MuRoom (Cerca Magnetics Ltd, UK)) equipped with active shielding using field nulling coils that reduced the remnant magnetic field <1 nT (21). Digitization of OPMs’ localization/orientation and face/head points was based on 3D optical scans (Einscan (Shining 3D); Figure 1). The set-up was adapted from (9–11,14). Patients were in comfortable sitting position, eyes closed, encouraged to fall asleep in order to induce IEDs (22), but not sleep deprived. Three patients (patients 8, 9 and 10) underwent on-scalp MEG recording after their clinical cryogenic MEG recording (Triux, MEGIN, Finland; 204 planar gradiometers, 102 magnetometers; sampling rate, 1,000 Hz; band-pass filter, 0.1-300 Hz, supine position) performed in a lightweight MSR (Maxshield (MEGIN), set-up detailed in (23)) in the context of presurgical assessment of RFE (as described in (2)). Six other patients were proposed to undergo a cryogenic MEG in the sitting position after the on-scalp MEG recording, which could thus have impacted sleep occurrence/duration during the subsequent second recording. During cryogenic MEG, patients’ head position was continuously tracked using four head position indicator coils. These coils and ∼300 face/scalp points were digitized relative to anatomic fiducials using an electromagnetic tracker (Fastrack Polhemus, USA). Two patients (patients 1 and 6) did not undergo cryogenic MEG due to the unavailability of the device for technical reasons (Table 1). Structural 3D T1-wheighted brain magnetic resonance imaging (MRI) was available from the clinical brain MRI in 8 patients (24). Three patients were offered a new structural brain MRI if the previous one was ≥10 years old for adults or ≥2 years old for children.

**Figure 1.**
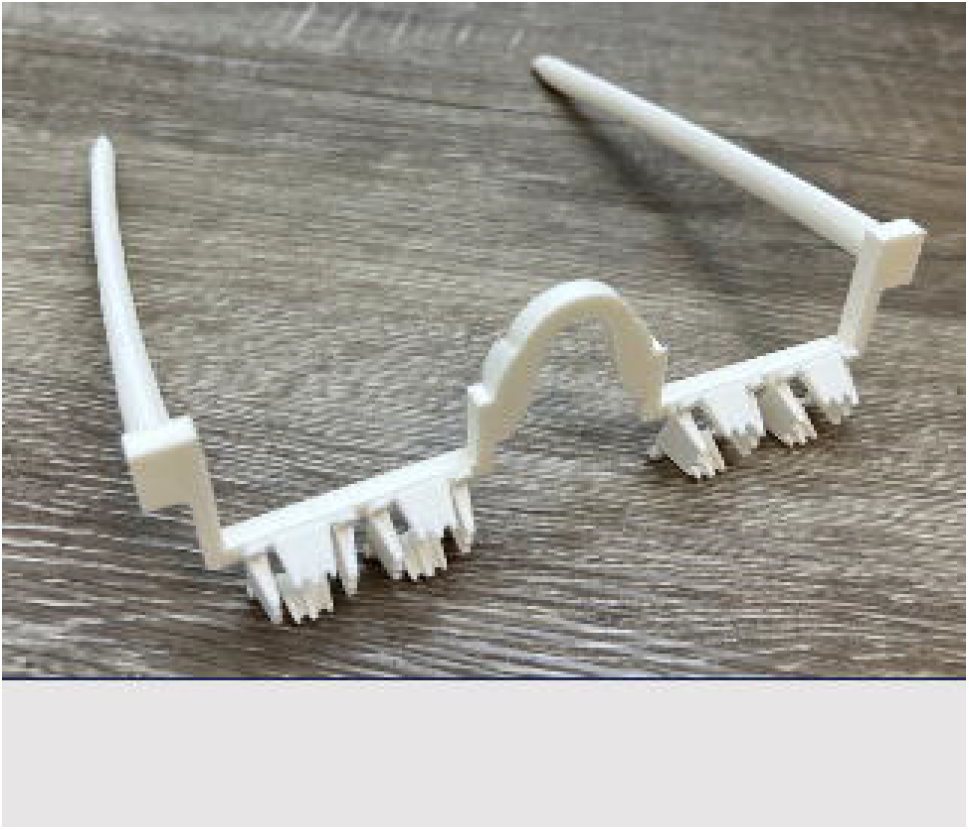
On-scalp MEG set-up including both scalp-OPMs and face-OPMs Top. Three-dimensional-printed glass-like structure that allows to place 4 OPMs on the face, right below the eyes (face-OPMs). Bottom. Healthy adult subject wearing flexible EEG-cap like (3D-printed holders sewn on the EEG-cap to place OPMs on the scalp (scalp-OPMs)) with 48 OPMs (28 Gen-2 OPMs (white upper cover) and 20 Gen-3 OPMs (blue upper cover)) and the glass-like structure with 4 Gen-3 OPMs. A cap with 2 reflexive markers is placed on top of each OPM to measure the localization and the orientation of the OPMs relative to the patients’ scalp using an optical scanner.

**Table 1.** Demographic and clinical data.

| N° | Patients (age, sex) | IED location, scalp EEG | IED location, cryogenic MEG (interval, on-scalp and cryogenic MEG) | Other clinical information |
| --- | --- | --- | --- | --- |
| 1 | Mean: 37y, 4F/7M | F7-T3 | MEG not available | / |
| 2 |  | F9-T9 | Left basal temporal (4 months) | / |
| 3 |  | T5-O1 | Left posterior temporal (same day) | / |
| 4 |  | F7-T3 | Left anterior temporal (same day) | / |
| 5 |  | F8-F4 | Right posterior temporal (same day) | / |
| 6 |  | F10-T10 | MEG not available | Right anterior temporal lobectomy to be performed after SEEG |
| 7 |  | F7-T3 | Left posterior temporal (same day) | / |
| 8 |  | F10-T10 | Right medial temporal (same day) | / |
| 9 |  | F8-T8 | Right basal temporal (same day) | SEEG to be performed |
| 10 |  | F8-T4 | Right medial temporal (15 months) | / |
| 11 |  | T4-C4 | Right temporal opercula (same day) | Seizure-free and IED-free after grade 1 ganglioglioma resection (at 1 year follow-up) |

Scalp EEG electrodes involved in IEDs is reported according to clinical EEG follow-up using the conventional 10-20 placement including inferior temporal electrodes. IED localization based on cryogenic MEG is reported similarly to the clinical MEG protocol. Patients 1 and 6 did not benefit from a cryogenic MEG recording. The delay with on-scalp MEG recording is notified under brackets. Some MEG recordings are performed in seating position to keep similar with on-scalp MEG recordings, after the on-scalp MEG recording when performed on the same day. MEG recordings performed in supine position were used for the clinical assessment of refractory focal epilepsy, before the on-scalp MEG recording when performed on the same day.
IED, interictal epileptiform discharge; MEG, magnetoencephalography; y, years; F, female; M, male; SEEG, stereo-electroencephalography.

## Data preprocessing and analyses

On-scalp MEG data were denoised using principal component analysis (removal of the first 15 components, optimal number based in visual inspection). Cryogenic MEG data used signal space separation with movement correction (Maxfilter, MEGIN, Finland) (25). MEG data were restricted to the 102 magnetometers for comparability (9). Both on-scalp and cryogenic MEG data were then band-pass filtered between 3-40 Hz.

Patients’ structural brain MRI’s were segmented using Freesurfer (26) and co-registered with on-scalp MEG digitalization using the face surface based on the method (27) implemented in the open-source Blender software, and with cryogenic MEG digitalization using anatomic fiducials and head surface points via the MRIlab software (MEGIN, Finland). Forward models were computed using the one-layer boundary element method (28).

IEDs were visually detected in both on-scalp and cryogenic MEG signals. Data were epoched around the peak of each detected IED and IEDs with similar spatio-temporal features were averaged. Source localization was performed on the averaged IED magnetic pattern at the peak time using custom-made dynamic statistical parametric mapping (29) with estimation of the noise covariance from baseline data (-100 to -50ms before the peak of IED) and regularization from the global SNR (30).

Peak IED amplitude and SNR (i.e., ratio of peak amplitude over baseline standard deviation) were assessed at the channel showing maximal IED response. Resulting values from on-scalp MEG and cryogenic MEG were compared using two-sided unpaired t tests (significance at p < 0.05) when a sufficient number (≥10) of IEDs was available in both data sets to allow proper statistical comparison and avoid small sample biases. The location of IED neural source obtained with on-scalp MEG was qualitatively described and its distance to that of cryogenic MEG IED was quantitatively compared.

With the aim to investigate the differences between ETLE and TLE, we compared the IED amplitude and SNR of patients from this study (TLE: n=11) to other patients from other studies that used the same set-up with flexible cap (9,10,31) (ETLE: n=7) using unpaired t-test. The low number of IEDs (<10) in some patients with TLE is acceptable for this analysis at the group level. We also performed a similar comparison for the distance between cryogenic and on-scalp MEG sources using unpaired t- test (TLE: n=9; ETLE: n=5 data extracted from (9)).

To investigate the added-value of face-OPMs, the distance between the neural sources reconstructed from both face- and scalp-OPMs vs. scalp-OPMs only was reported.

Changes in OPM proximity to brain source brought by face-OPMs was also assessed by reporting the distance between the IED source localization (based on both face- and scalp-OPMs) and either face- or scalp-OPMs for each patient and comparing them with two-sided paired t-test (p < 0.05). A larger distance for face-OPMs compared to scalp-OPMs would likely indicate lesser sensitivity of face-OPMs to IED activity. We further assessed whether face- and scalp-OPMs sense similar brain activity using canonical correlation, i.e., a multivariate measure of temporal correlation between the set of signals (averaged IED time course) recorded by face-OPMs and the set of signals recorded by scalp-OPMs. For each patient, statistical significance of this canonical correlation was established non-parametrically by generating 10,000 sets of surrogate face- and scalp-OPMs rendered independent by global randomization of the Fourier phase of each set of signals (hence preserving the correlation structure within each set) and estimating the p value as the fraction of surrogates whose canonical correlation exceeded the original estimate.

## Results

Eleven patients (4F/7M) with TLE were included. Demographic and clinical data are detailed in Table 1. Depending on technical availability, from 43 to 53 OPMs were used, leading to 98 to 127 usable channels/patient (Table 2). IEDs were detected with on-scalp MEG in all patients (mean number of IEDs: 13 IEDs/patient, mean amplitude of IEDs: 3.3 pT, mean SNR of IEDs: 9.4) and localized in the temporal lobe (anterior, n=2; medial, n=2; basal, n=3; lateral, n=2; posterior, n=2; Table 2). Four patients had a sufficient number (≥ 10) of IEDs to have amplitude/SNR statistically compared with cryogenic MEG IEDs, but one of them did not undergo cryogenic MEG due to a technical issue. Among the three other patients, Patient 3 had higher amplitude and SNR of IEDs with on-scalp MEG, Patient 9 had higher amplitude but similar SNR, and Patient 10 had similar amplitude and SNR with both modalities (Table 2). On-scalp and cryogenic MEG sources were separated by 6-16 mm (Figure 2). Patient 6 underwent stereo-electroencephalography (SEEG), which confirmed the localization of the irritative zone highlighted by on-scalp MEG. Patient 11 underwent resective surgery with spatial concordance between the irritative zone highlighted by on-scalp MEG and the resection cavity (Figure 3). In 10/11 patients, face-OPMs did not substantially modify the IED source localization compared with scalp-OPMs only (mean distance between sources for patients 2 to 11: 3.5 mm, range: 0-11 mm; Table 3). In Patient 1, IEDs were mostly observed on left face-OPMs compared with scalp-OPMs, which substantially impacted source reconstruction (Table 3 & Figure 4). Overall, face-OPMs were significantly more distant from IED neural sources than scalp-OPMs (Table 3, p = 3.26x10^-7^). In 9 of these 10 patients, a significant correlation was found between face- and scalp-OPM signals suggesting that face-OPMs were recording corresponding brain activity (Table 3 & Figure 5).

**Figure 2.**
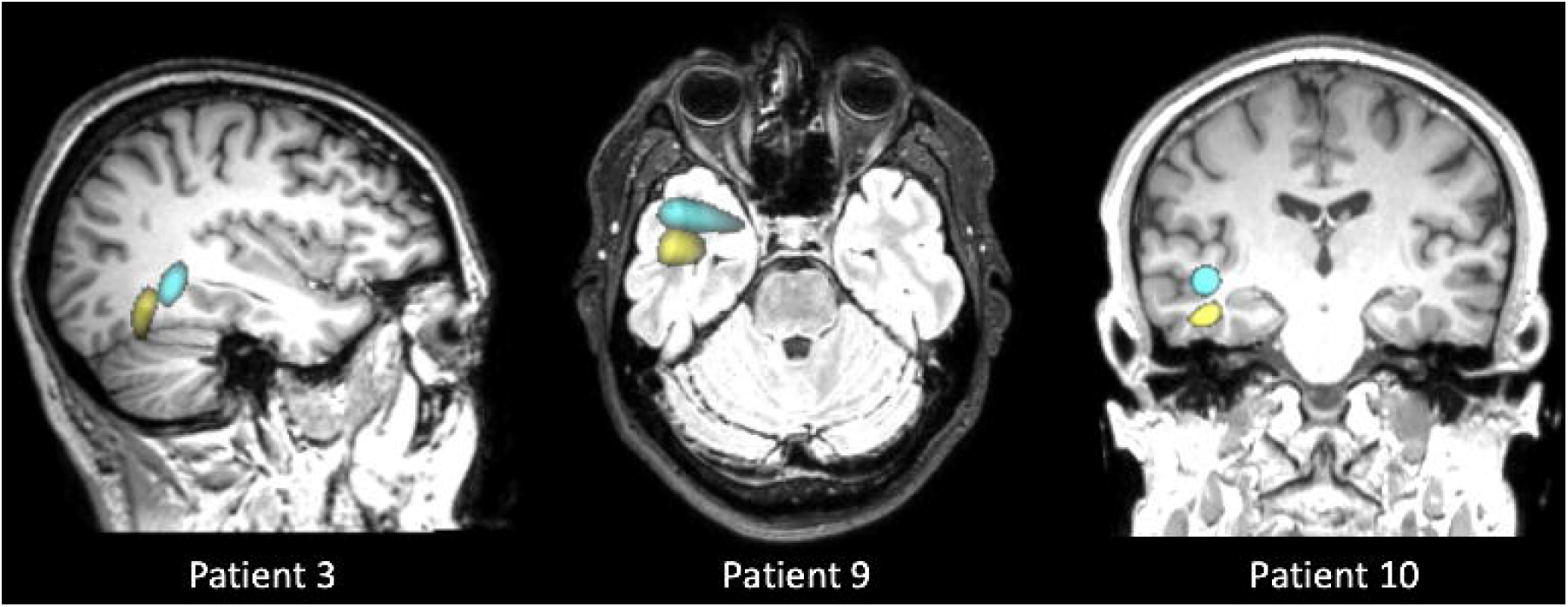
On-scalp vs. cryogenic MEG On-scalp (yellow) vs. cryogenic MEG (blue) neural IED source localization superimposed on individual T1-weighted brain MRI. Left. Sagittal slice through the left hemisphere of Patient 3. Middle. Axial slice of Patient 9. Right. Coronal slice of Patient 10. MRIs are shown in radiological convention.

**Figure 3.**
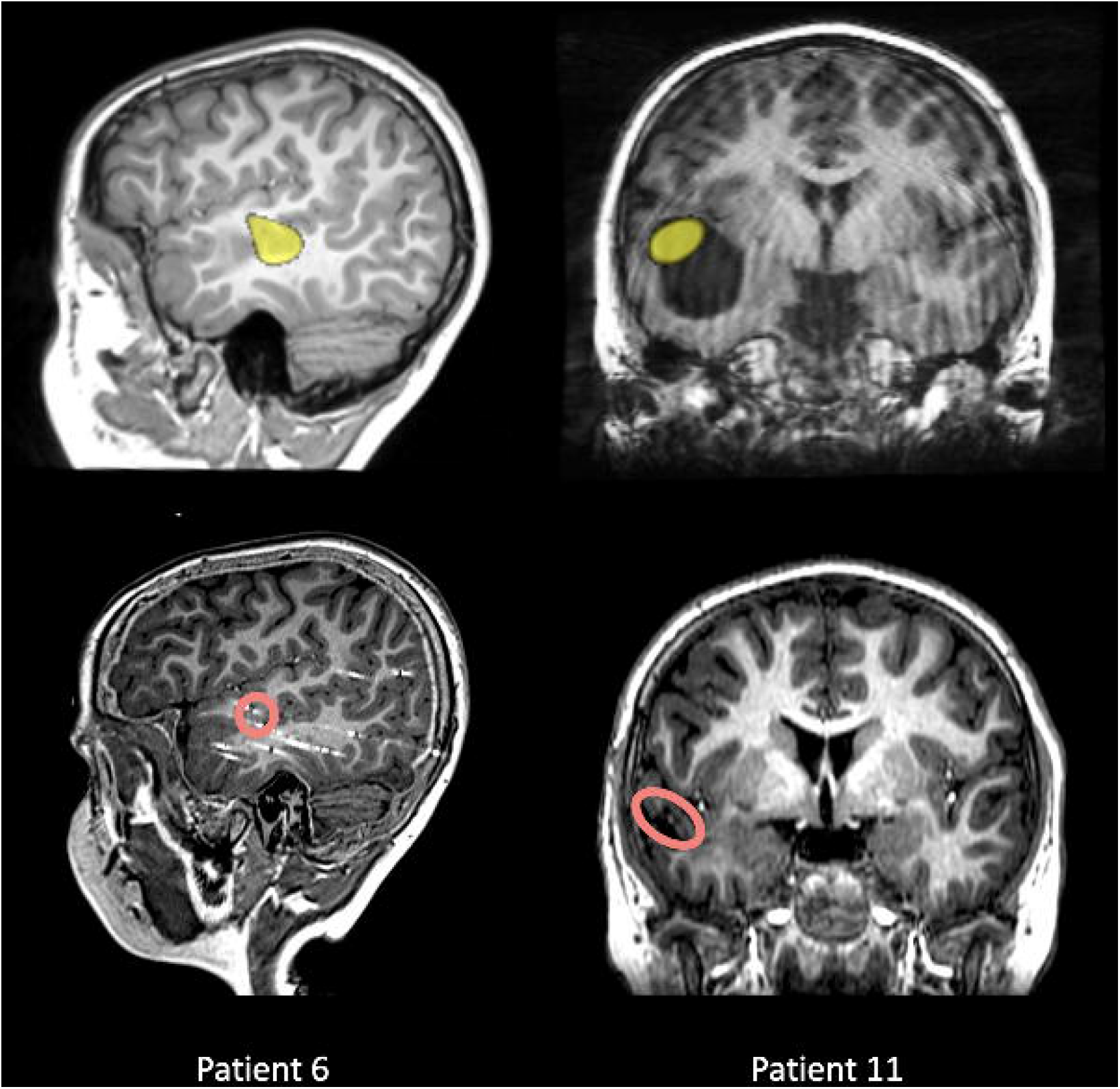
Comparison of on-scalp MEG with invasive gold-standards Left. On-scalp MEG source localization (yellow) displayed on a sagittal slice of T1-wheighted brain MRI through the right hemisphere of patient 6 (top) and depth electrodes implanted in the right hemisphere during the SEEG subsequent to the on-scalp MEG recording on a co-registered pre- implantation T1-wheighted brain MRI and post-implantation brain computed tomography (bottom). The two corresponding depth electrodes (red circle) were involved in the irritative zone during SEEG recording, which validates the on-scalp MEG IED neural source. Right. On-scalp MEG source localization (yellow) displayed on a coronal slice of T1-wheighted brain MRI of patient 11 suffering from grade 1 ganglioglioma (top) and post-resection coronal slice of T1-wheighted brain MRI (bottom). The on-scalp MEG source was involved in the lesion and included in the area of resection, leading to seizure-freedom (Engel class IA) and IED-freedom at one-year post-surgery. Pre- and post- implantation look different due to mass effect on the pre-implantation imaging and brain shift on the post-implantation imaging. MRIs are shown in radiological convention.

**Figure 4.**
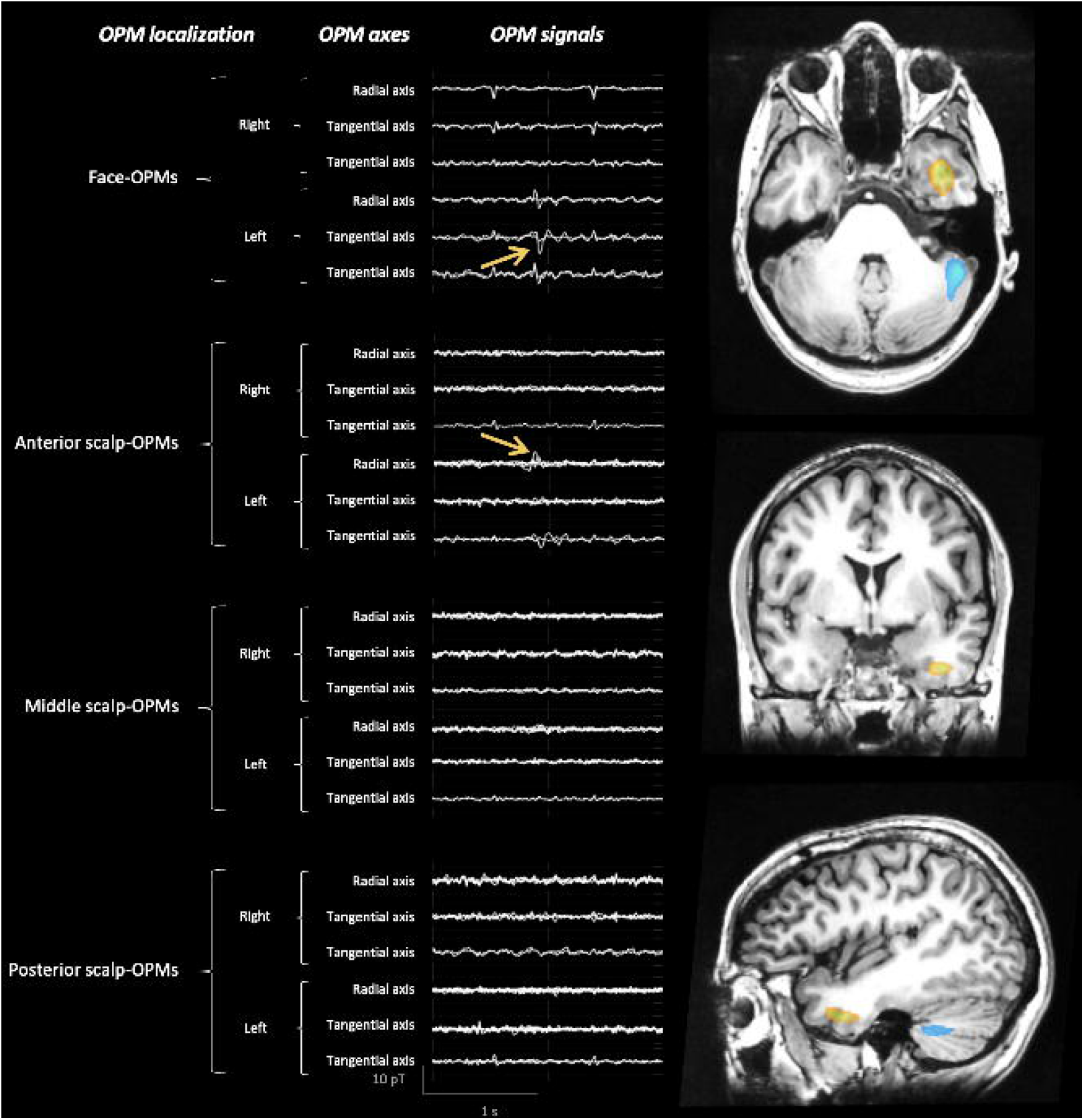
On-scalp MEG signals and IED neural sources with/without face-OPMs. Left. On-scalp MEG signals of Patient 1 showing an anterior temporal IED best sensed by left face- OPMs compared with left scalp-OPMs (yellow arrows). Right. On-scalp MEG IED source localizations inferred from both face-OPMs and scalp-OPMs (yellow) and from scalp-OPMs only (blue) displayed on T1-wheighted brain MRI (top, axial slice; middle, coronal slice; bottom, sagittal slice through the left hemisphere) in radiological convention.

**Figure 5.**
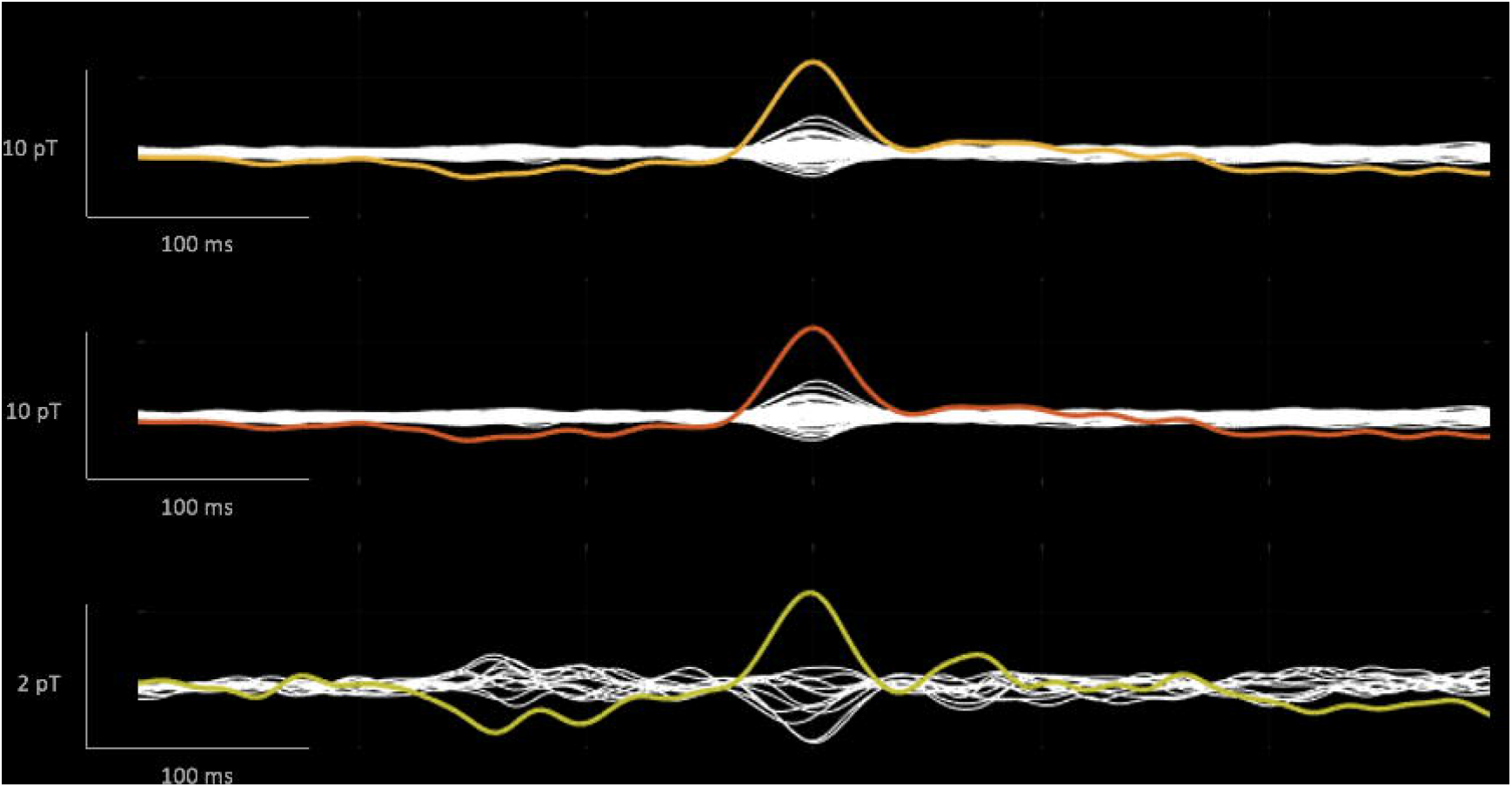
Correlation between scalp-OPMs and face-OPMs. On-scalp MEG signals averaged at the peak of the IEDs of Patient 9. Top. Signals including all OPMs and their principal component (yellow). Middle. Signals including scalp-OPMs and their principal component (red). Bottom. Signals including face-OPMs and their principal component (green).

**Table 2.** On-scalp MEG based on OPMs results and comparison with cryogenic MEG.

| N° | OPMs (channels) | IEDs frequency | IEDs localization | IEDs amplitude |  | IEDs SNR |  | Distance between on-scalp and cryogenic MEG |
| --- | --- | --- | --- | --- | --- | --- | --- | --- |
|  |  |  |  | OPMs | Cryogenic (p-value) | OPMs | Cryogenic (p-value) |  |
| 1 | 48 (113) | Rare | Left temporal pole | $7.2 \pm 1.0$ pT | / | $11.5 \pm 2.6$ | / | / |
| 2 | 53 (127) | Rare | Left middle T3-T4 sulcus | $3.1 \pm 1.2$ pT | $1.3 \pm 0.03$ pT | $7.9 \pm 1.6$ | $3.8 \pm 0.5$ | 16 mm |
| 3 | 44 (102) | Frequent | Left posterior T4 gyrus | $4.0 \pm 0.4$ pT | $2.2 \pm 0.3$ pT ($6.0 \times 10^{-4}$) | $11.5 \pm 2.0$ | $5.4 \pm 0.9$ ($7.0 \times 10^{-3}$) | 9 mm |
| 4 | 43 (98) | Few | Left anterior T1-T2 sulcus | $2.7 \pm 0.3$ pT | $0.3 \pm 0.07$ pT | $11.6 \pm 2.1$ | $2.7 \pm 1.4$ | 11 mm |
| 5 | 53 (127) | Few | Right posterior T3 gyrus | $2.4 \pm 0.4$ pT | $2.3 \pm 0.2$ pT | $3.4 \pm 0.5$ | $5.5 \pm 1.2$ | 15 mm |
| 6 | 53 (127) | Frequent | Right middle T1-T2 sulcus | $2.9 \pm 0.3$ pT | / | $7.1 \pm 1.0$ | / | / |
| 7 | 51 (124) | Few | Left posterior T2 gyrus | $2.1 \pm 0.3$ pT | $1.2 \pm 0.4$ pT | $6.0 \pm 1.6$ | $3.9 \pm 0.7$ | 14 mm |
| 8 | 52 (124) | Rare | Right peri-amygdala and peri-hippocampal | $4.0 \pm 2.0$ pT | $1.5 \pm 0.3$ pT | $21.5 \pm 16.3$ | $22.5 \pm 10.7$ | 6 mm |
| 9 | 44 (105) | Frequent | Right temporal pole | $2.4 \pm 0.2$ pT | $1.0 \pm 0.03$ pT ($6.7 \times 10^{-7}$) | $9.6 \pm 1.2$ | $13.5 \pm 3.6$ (0.32) | 8 mm |
| 10 | 48 (116) | Abundant | Right peri-hippocampal | $1.4 \pm 1.7$ pT | $1.2 \pm 1.7$ pT (0.31) | $3.6 \pm 0.6$ | $4.4 \pm 0.5$ (0.26) | 11 mm |
| | | | Right posterior T1 | $1.9 \pm 0.1$ pT | / | $7.5 \pm 1.2$ | / | / |
| 11 | 43 (102) | Rare | Right temporal opercula | $6.0 \pm 1.7$ pT | $1.7 \pm 0.08$ pT | $11.2 \pm 6.2$ | $9.0 \pm 3.3$ | 7 mm |

Legend for the IEDs frequency: rare, <5; few, <10; frequent, $\geq 10$ ; abundant, >50 IEDs during 1-hour on-scalp MEG recording. Amplitudes and SNR are reported as mean and standard error on the mean. MEG, magnetoencephalography; OPM, optically pumped magnetometer; IED, interictal epileptiform discharge; SNR, signal-to-noise ratio.

**Table 3.**
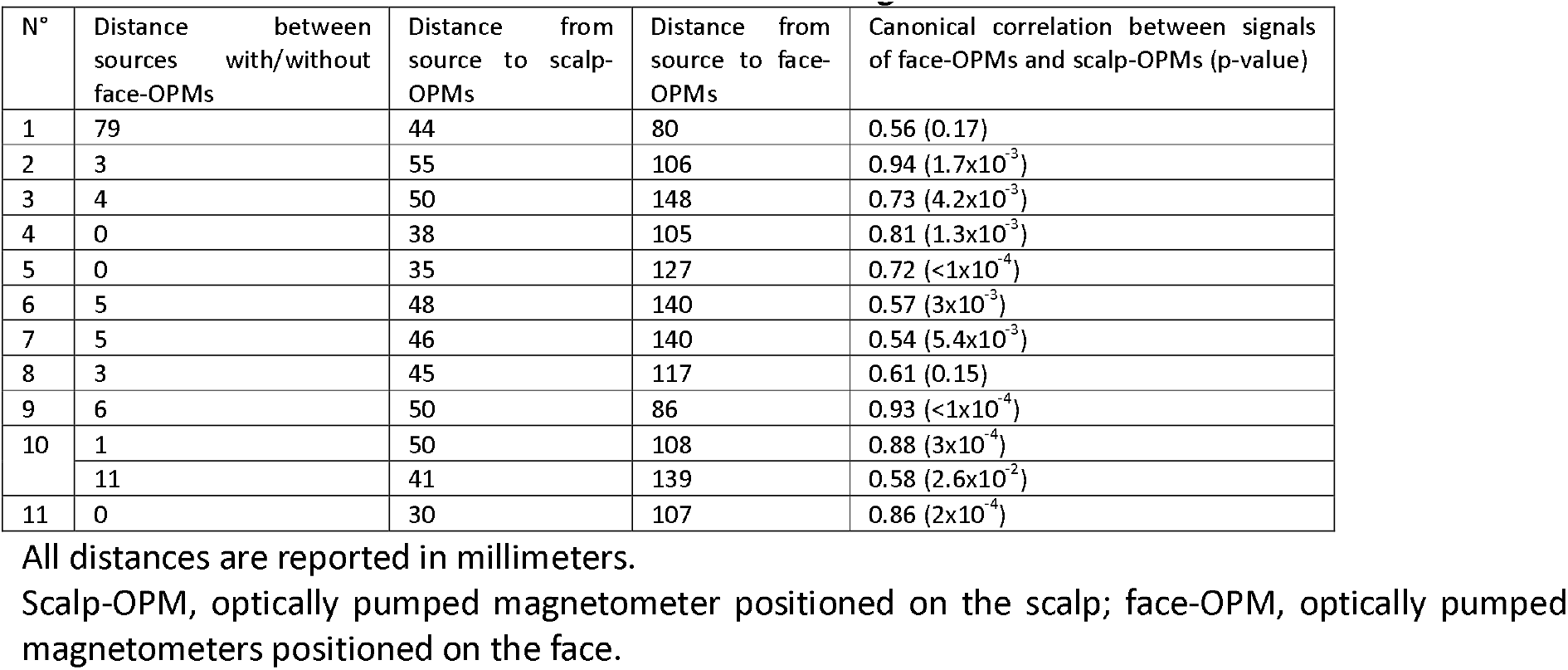
Contribution of face-OPMs to recorded brain signals.

The reported amplitudes of IEDs (mean: 3.3 pT, range: 1.4-7.2 pT) detected by on-scalp MEG tended to be smaller than ETLE IEDs (mean: 6.0 pT, range: 2.5-9.9 pT, p = 0.05), with significantly lower SNR (TLE, mean: 9.4, range: 3.4-21.5; ETLE, mean: 14.3, range: 11.1-21.3; p = 0.02). The distance between cryogenic and on-scalp MEG sources did not differ significantly between TLE and ETLE (TLE: 6 to 16 mm, ETLE: 4 to 16 mm, p = 0.23).

## Discussion

This study demonstrates in a group of 11 patients with TLE that on-scalp MEG can accurately detect and localize temporal IEDs. It also shows that additional face-OPMs may play a key role in some patient for the detection of antero-mesial temporal IEDs.

### Yield of on-scalp MEG in TLE

To the best of our knowledge, this study reports the most important population of epileptic patients investigated using on-scalp MEG to date. The lower SNR and tendency for a lower amplitude in TLE compared with ETLE is probably due to the deep location and partial coverage of the irritative zone in some of the included patients (Patients 1, 8-10; Table 2). Two previous case-studies also investigated patients with TLE using on-scalp MEG (12,14). Amplitudes and SNR of epileptiform discharges cannot be compared with our data due to the use of rigid helmets in both of them, the study of temporal ictal discharges on the one hand (12), and the presence of simultaneous SEEG electrodes on the other hand (14). The distance between cryogenic and on-scalp MEG neural sources does not significantly differ between TLE patients and ETLE patients, which confirms the ability of on-scalp MEG to discriminate a large variety (medial, anterior, basal, lateral, posterior) of temporal sources.

Among the 11 included patients, 3 had a sufficient number of IEDs detected with on-scalp and cryogenic MEG to allow proper comparison between the two modalities. In contrast with our previous studies, this comparison was based on a similar number of magnetic field sensors (i.e., 102 cryogenic magnetometers vs. 102-116 OPM channels, p = 0.31). It showed that IED amplitude and SNR were either similar or higher with on-scalp MEG than with cryogenic MEG, similarly to previous results in children with ETLE (9). First, Patient 3 had a higher IED amplitude and higher IED SNR with on-scalp MEG than cryogenic MEG, which is comparable to our previous study in school-aged children (9) despite higher head circumference and lower IED frequency. Second, Patient 9 had a higher IED amplitude with on-scalp MEG but similar SNR using both on-scalp and cryogenic MEG. The cryogenic MEG was recorded in the supine position before (in the morning) on-scalp MEG recording in the sitting position (in the afternoon, same day). This could have negatively impacted sleep quality during on-scalp MEG due to IEDs (32) and/or noise modifications due to movement differences between sleep and wakefulness. Third, Patient 10 had similar IED amplitude/SNR, but with an interval between on-scalp and cryogenic MEG of 15 months, which renders difficult the interpretation of this comparison. Similarly, the detection of a second irritative zone in that patient is difficult to interpret because of this delay. Despite those limitations, these comparative results are of critical importance as they suggest, as shown previously in other cases of ETLE (9), that on-scalp MEG is non-inferior, and in some cases superior, to cryogenic MEG for the amplitude/SNR of detected IEDs.

The neural sources of IEDs reconstructed based on on-scalp and cryogenic MEG were separated by ∼10 mm, which is slightly higher than the recognized spatial resolution of cryogenic MEG (∼5 mm (16)) but close to what has been previously showed in children with ETLE (9). Possible hypotheses for this increased distance could be the higher spread of IEDs during sleep (33) or the distinct spatial pattern depending on the vigilance state during recordings (34). These hypotheses are supported by the fact that patients reported difficulties to fall asleep twice in a row. It could also be due to the fact that recordings were not done simultaneously and that they thus did not sample similar IEDs from the irritative zones. It could also be due to inaccuracies in the estimation of OPM location/orientation/calibration (35), which could be alleviated by the development of automatic OPM calibration/co-registration, or a lower spatial resolution for on-scalp MEG because of lesser spatial sampling and less efficient noise cancellation (36).

Of note, similar findings (amplitude and SNR levels, distance between reconstructed neural sources) between on-scalp and cryogenic MEG were obtained in the other patients (Patients 2, 4, 5, 7, 8, 11) with a low (<10) IED number (no formal statistical comparison due to the low IED number).

In two patients, we were able to compare the results of on-scalp MEG with “gold-standard” methods of localization of the epileptogenic zone. The epileptogenic zone has two possible definitions: the smallest cortical area to be resected to make the patient seizure-free (37) that can be approximated by the resected area, or the area where seizures started and primary organized (38) based on SEEG data. In Patient 11, the surgical resection cavity comprised the neural source of IEDs highlighted by on-scalp MEG (Figure 3). The patient being seizure-free (Engel class IA) and IED-free at 1-year post-surgery, this finding validates our on-scalp MEG findings. In Patient 6, SEEG demonstrated that the irritative zone involved the plots of depth electrodes (red circles on Figure 3) that were located at the on-scalp MEG source (Figure 3).

### Added value of face-OPMs

A clear advantage of on-scalp MEG compared with cryogenic MEG is that OPMs can be placed at various locations on the head to improve the sensitivity of OPM arrays to specific neural sources (39,40). We previously demonstrated using a low number of tri-axial OPMs that different OPM position at/around the irritative zone can lead to differences (involved axes, amplitude, SNR) in IED detection between OPMs (31). Our innovative placement of face-OPMs on a 3D-printed glass-like structure makes it more comfortable and adapted to clinical applications than mouth-OPM (17), and non-invasive compared with sphenoidal electrodes (18). Increasing the density of sensors around the area of interest is known to increase the sensitivity of the technique (41), and so does increasing sensor coverage (17,20). Face-OPMs thus represent a unique opportunity to increase the yield of on-scalp MEG in anterior TLE due to the vicinity of sensors with the investigated brain area or medial TLE due to the lead field pattern of hippocampal activity on the palate (17). The added-value of face- OPMs is less clear in case of posterior neocortical TLE, although their signals were correlated with scalp-OPM IEDs. While face-OPMs are more distant from the brain than scalp-OPMs, they are still able to capture brain signals as shown by this correlation between their signals (Table 3 & Figure 5) and thus effectively increased OPM spatial sampling. Of note, the correlation was not found significant in patient 1 and patient 8, which does not necessarily mean that brain activity was undetected by face-OPMs, but that different cerebral activity could be detected by face-OPMs. The only way to confirm this hypothesis with certainty would be to compare face-OPMs and scalp-OPMs with simultaneous intracranial EEG.

Patient 1 particularly benefited from face-OPMs for IED detection and source reconstruction (Figure 4). Indeed, IEDs were mainly captured by left face-OPMs compared with scalp-OPMs leading to completely different source reconstruction results when including face-OPMs or not. Moreover, the neural source location obtained when including face-OPMs was in agreement with results of previous scalp EEG (i.e., electrodes F7-T3). While benefiting from a higher SNR than EEG for temporal pole sources, cryogenic MEG demonstrates lower SNR at this cortical area compared with other neocortical sources (42). The difference in IED detection between face- vs. scalp-OPM in this patient could be related to the localization of sources (i.e., temporal pole) or to the orientation of sources (that is not assessed using distributed source reconstruction). The orientation of cortical sources could be partly related to their localization, brain morphology and cortical folding. As the cortical surface is not perfectly spherical, the use of a realistic head model is known to be useful for orientation sources to which the MEG is less sensitive (43) and the addition of face-OPMs on glass-like structure could also help detecting these sources. Patient 9 had also a temporopolar source with no real added-value of face-OPMs, that could be due to the orientation of the source or to the cortical extent of the IED activity. Indeed, the minimal cortical surface needed to generate a magnetic field detectable on scalp depends on the depth of the source (44) and propagation of the activity to a larger cortical area renders detection by MEG more probable. Cryogenic MEG is completely blind to only 5% of the neocortical activity that originates from sources with orientation that deviates from less than 15° compared to the head radial direction (45). Still, the increased spatial coverage of the temporal lobe may be of interest in patients with TLE, even if no substantial difference is found between on-scalp MEG with or without face-OPMs. The maximal yield of face-OPMs could thus be reached in patients with suspected medial/anterior TLE.

### Limitations

This study is first limited by the absence of simultaneous gold-standard (i.e., SEEG) or conventional (i.e., scalp EEG) recordings to assess that similar IEDs were recorded between both on-scalp and cryogenic MEG (46) and to be used as a ground-truth confirmation (for SEEG only) of the source localization (14). Second, the absence of cryogenic MEG, that is the most similar type of recording, in patients 1 and 6 and the delay between on-scalp and cryogenic MEG in patients 2 and 10 makes the comparison between both methods impossible or difficult in 40% of the patients. Additionally, the low number of detected IEDs in seven patients impedes the quantitative comparison in these patients due to the risk of small samples biases.

### Conclusion and perspectives

On-scalp MEG is able to detect and localize IEDs originating from medial and neocortical temporal areas. It is thus a promising tool to investigate patients with refractory TLE, which provides similar or superior findings compared with cryogenic MEG. The use of face-OPMs proved useful to detect anterior temporal epileptiform discharges. This study thus paves the way for further application of on-scalp MEG in RFE and in other neurological disorders involving the temporal lobe such as, e.g., Alzheimer’s disease (47).

## Data Availability

Data are available upon reasonable request to the corresponding author and after approval by institutional (Hopital universitaire de Bruxelles & Universite libre de Bruxelles) authorities.

## Authors contribution

O.F. designed and conceptualized the study. O.F., C.D., E.R., N.G., W.V.P., A.A., O.B., E.C., X.D.T. contributed to the patients’ recruitment. O.F., V.W., N.H., M.B., M.F., P.C., X.D.T. contributed to the set- up of acquisition. O.F. performed acquisitions. O.F., V.W. contributed to the analysis of data. O.F. and X.D.T. interpretated the data. X.D.T. supervised the study. X.D.T. obtained funding. O.F., V.W., C.D., E.R., N.G., W.V.P., A.A., O.B., E.C., N.H., M.B., M.F., P.C., X.D.T. contributed to draft or review the manuscript.

## Acknowledgments

O.F. is supported by the Fonds pour la formation à la recherche dans l’industrie et l’agriculture (FRIA, Fonds de la Recherche Scientifique (FRS-FNRS), Brussels, Belgium). P.C. was supported by the Fonds Erasme (Convention « Alzheimer », Brussels, Belgium). P.C. is supported by a research grant of INNOVIRIS (Brussels, Belgium; “DetectDem” research grant). M.F. is supported by the Fonds Erasme (Convention « Alzheimer », Brussels, Belgium). X.D.T. is Clinical Researcher at the FRS-FNRS. The other authors received no additional funding.

The on-scalp MEG project at the Hôpital Universitaire de Bruxelles and Université libre de Bruxelles is financially supported by the Fonds Erasme (Projet de Recherche Clinique et Convention « Les Voies du Savoir ») and by the FRS-FNRS (Crédit de Recherche: J.0043.20F, Crédit Équipement: U.N013.21F).

